# Data-driven identification of potentially successful intervention implementations: a proof of concept using five years of opioid prescribing data from over 7000 practices in England

**DOI:** 10.1101/2023.06.28.23291704

**Authors:** Lisa E. M. Hopcroft, Helen J. Curtis, Richard Croker, Felix Pretis, Peter Inglesby, Dave Evans, Seb Bacon, Ben Goldacre, Alex J. Walker, Brian MacKenna

**Affiliations:** Bennett Institute for Applied Data Science, Nuffield Department of Primary Care Health Sciences, University of Oxford, OX2 6GG, UK; Department of Economics, University of Victoria, Victoria, BC, V8P 5C2, Canada

**Keywords:** Electronic health records, primary care, general practice, opioid analgesics, data science, implementation science

## Abstract

**Background:** We have previously demonstrated that opioid prescribing increased by 127% between 1998 and 2016. New policies aimed at tackling this increasing trend have been recommended by public health bodies and there is some evidence that progress is being made. We sought to extend our previous work and develop an unbiased, data-driven approach to identify general practices and clinical commissioning groups (CCGs) whose prescribing data suggest that interventions to reduce the prescribing of opioids may have been successfully implemented.

**Methods:** We analysed five years of prescribing data for three opioid prescribing measures: one capturing total opioid prescribing and two capturing regular prescribing of high dose opioids. Using a data-driven approach, we applied a modified version of our change detection Python library to identify changes in these measures over time, consistent with the successful implementation of an intervention. This analysis was carried out for general practices and CCGs, and organisations were ranked according to the change in prescribing rate.

**Results:** We present data for the three CCGs and practices demonstrating the biggest reduction in opioid prescribing across the three opioid prescribing measures. We observed a 40% drop in the regular prescribing of high dose opioids (measured as a percentage of regular opioids) in the highest ranked CCG (North Tyneside); a 99% drop in this same measure was found in several practices. Decile plots demonstrate that CCGs exhibiting large reductions in opioid prescribing do so via slow and gradual reductions over a long period of time (typically over two years); in contrast, practices exhibiting large reductions do so rapidly over a much shorter period of time.

**Conclusions:** By applying one of our existing analysis tools to a national dataset, we were able to rank NHS organisations by reduction in opioid prescribing rates. Highly ranked organisations are candidates for further qualitative research into intervention design and implementation.

**Contributions to the literature:**

- Demonstrating that a data-driven approach can identify and quantify changes in important clinical measures in publicly available NHS data
- Identifying changes in this way allows the unbiased identification of candidates for further qualitative research into intervention design and implementation
- Large reductions observed at the CCG level (which are more robust to local circumstances) demonstrate that it is possible to reduce opioid prescribing and that continued and wider success in reducing opioid prescribing is dependent, at least in part, to closing an implementation gap

## Background

The prescription of opioids is common and appropriate in the management of acute pain but their efficacy with regards to chronic pain is not supported by empirical evidence (1). Long term use of opioids has been shown to be associated with accumulating risk of dependence and overdose (2). Continually rising rates of opioid prescription, particularly in England and Wales (3–5), prompted the publication of new guidance in 2010 (6) advocating for a cautious approach in the long term prescribing of opioids (7), and opioids have been a specific priority for governmental advisory groups (8). In 2019, Public Health England (PHE) published the Prescribed Medicines Review which aimed to “identify the scale, distribution and causes of prescription drug dependence, and what might be done to address it” (9). This review included data from the NHS Business Services Authority’s (NHSBSA) primary care prescription dataset, which suggested some progress had been made in reducing opioid prescribing, with a small but consistent fall in rates between 2015 and 2018. However, there was also evidence that opioid prescribing remains a persistent public health problem in England, with higher rates of prescription in areas of higher deprivation and evidence that long term prescribing was associated with opioid overdose and dependence. The first recommendation of this report was “increasing the availability and use of data on the prescribing of medicines that can cause dependence or withdrawal to support greater transparency and accountability and help ensure practice is consistent and in line with guidance” (9).

Our group produces <u>OpenPrescribing.net</u>, which allows open access to the same NHSBSA primary care prescription dataset as used in the PHE review. It is a free and widely used tool with 20,000 unique users per month, where anyone can explore the prescriptions dispensed at any practice in England and monitor prescribing patterns down to the level of individual brands, formulations and doses.

In OpenPrescribing, we perform automated analyses to generate monthly reports covering 80 measures of prescribing safety, effectiveness and cost for multiple administrative levels of NHS England, including all general practices and clinical commissioning groups (CCGs), with functionality to visualise changes over time. Several measures have been developed to capture trends and variation in opioid prescribing (10) (Box 1 describes how to access opioid prescribing data for NHS organisations of interest). This window onto national opioid prescribing data presents an opportunity to identify changes—both increases and decreases—in prescribing that could inform NHS decision making and policy.

It is our experience that best practice is typically defined by organisations identifying *themselves* as having improved, following implementation and internal assessment of interventions. We are seeking to pursue an alternative, data-driven, unbiased approach which instead exploits the national prescribing dataset to identify prescribing patterns that may be representative of best practice (i.e., where we can identify a significant reduction in opioid prescribing).

We set out to support the first recommendation of the PHE review by applying our change detection algorithm (11) to identify patterns indicative of sustained and significant reduction that may help identify best practice with regards to opioid prescribing policy. Applying this to multiple administrative levels of the NHS, we identified specific practices and CCGs whose prescribing data indicates that they have successfully implemented an intervention to reduce prescribing of opioids, and are important targets for further qualitative research.

### BOX 1 Accessing opioid prescribing data for specific organisations via OpenPrescribing

Opioid prescribing data are publicly available on <u>OpenPrescribing.net</u> for all current practices (providers of primary care) and Clinical Commissioning Groups (CCGs, an NHS administrative grouping of practices at the time of the study) in England, for the last five years.

To find an organisation’s data for any of the opioid measures described in this manuscript, navigate to the specific measure’s landing page (12–14). Decile plots for all current CCGs are available on this page; specific CCGs can be identified via a text search in your browser. To view similar decile plots for a practice, click the “*Split the measure into chargers for individual practice*” link under the parent CCG. All opioid measures for a single organisation can be viewed as described in <u>this Youtube tutorial</u>.

Alternatively, summary results can be obtained for more than one organisation at once by selecting “View this measure on the analyse page” (under “Explore”) on the measure’s landing page (12–14); this will launch a new analysis, pre-loaded with the relevant drugs or BNF sections. Any number of organisations (CCGs or practices) can then be selected by typing code or text into the “highlighting” box; clicking on the “Show me the data!” button will launch this analysis and display the results as a histogram, time series or as a choropleth map. All plots and raw data are available for download.

## Methods

### Study design

We conducted a retrospective database study using general practice primary care electronic health record (EHR) data from all GP practices in England.

### Data source

We extracted data from the OpenPrescribing.net database. This imports openly accessible prescribing data from the large monthly files published by the NHS Business Services Authority, which contain data on cost and items prescribed for each month, for every typical general practice and CCG in England since mid-2010 (15). We extracted data up to May 2019. We note that CCGs were replaced by ICBs (integrated care boards) as of 1st July 2022. We have retained results by CCGs as this was an active administrative unit of the NHS in England during the study period. The monthly prescribing datasets contain one row for each different medication and dose, in each prescribing organisation in NHS primary care in England, describing the number of items (i.e., prescriptions issued) and the total cost. These data are sourced from community pharmacy claims data and, therefore, contain all items that were dispensed. We extracted all available data for typical general practices, excluding other organisations such as prisons and hospitals, according to the NHS Digital dataset of practice characteristics and excluded practices that had not prescribed at least one item per measure. The numbers of patients registered at each practice were obtained from NHS Digital. Practices were excluded for any month in which they had no registered patients or no prescribing.

### Study measures

Three measures were used in this study to capture various aspects of opioid prescribing. The first (“*Total Oral Morphine Equivalence per 1000 Patients”*) expresses the oral morphine equivalence (OME) of *all* opioid prescriptions per 1000 patients (12). The second and third look to capture information about regularly prescribed opioids: those used on a regular basis to control pain rather than preparations used for breakthrough pain or opioid injections. Of the regularly prescribed opioids, high dose opioids were defined as those with _≥_120mg OME per day. The ^“^*High dose opioids as percentage regular opioids*^”^ measure captures the number of prescriptions of these high dose, regularly prescribed opioids as a percentage of all long-acting opioids (13); the ^“^*High dose opioid items per 1000 patients*^”^, captures the same number of high dose, long-acting opioids but expresses this per 1000 patients (14). For all measures, higher values represent higher rates of opioid prescription.

In England, an individual will be registered at one General Practice (GP or practice); and each practice will belong to a parent Clinical Commissioning Group (CCG). These organisations and the relationship between them can change over time (for example, a practice may be reassigned to a different CCG; a CCG may be renamed or replaced; or a practice may close). In our results, we report results for any practice or CCG that existed during the study period, acknowledging that some of these no longer exist.

Monthly values for each measure were calculated for every practice and CCG between December 2014 and November 2019 (this study period was chosen so as to assess prescribing rates over a reasonable period of time, without being affected by the COVID-19 pandemic). The monthly data were summarised as deciles and presented as decile charts across all practices or CCGs each month.

### Statistical methods

For this study, we used our innovative change detection Python library (<u>available via the Python Package Index</u>), which is an automated method of detecting change in time-series data. This algorithm was originally developed to determine how clinicians vary in their response to new guidance on existing or new interventions, by measuring the timing and magnitude of change in the relevant organisations; it is able to identify both steep, sudden changes and more gradual, smooth transitions over multiple months. The full methods are described elsewhere (11) and the code is available for anyone to use as a single command with our open Python library (16).

Data for each of the three measures were analysed for all 191 CCGs and 7458 practices. The time series for each organisation was analysed using our change detection algorithm (using the default parameters) to identify the location and magnitude of significant *reductions* in the measure (substantial *increases* were filtered out as they are not relevant to the research question). These results were then filtered to remove (i) 678 closed or dormant practices; and (ii) a further 237 practices with a list size of less than 2000 (this latter group was excluded to avoid analyses of time series with a high level of noise due to low prescribing volume); this process left 6543 practices to be subject to further analysis. Amongst the organisations where our code detected a substantial reduction, we selected those whose starting level immediately before the reduction was in the top 20% of all peer organisations; this was to remove any organisations with consistently low prescribing from our results. We then ranked practices and CCGs by the total measured change (the % reduction between the pre-drop value and the end-drop value) to identify which organisations exhibited the most substantial reductions.

The decile plots provided show an individual organisation’s prescribing rates across the period (thick red line), in the context of all peer organisations (summarised using deciles, as blue lines).

### Software and reproducibility

Data management and analysis was carried out using Python 3.8 and Google BigQuery, with analysis carried out using Python. Our change detection library (<u>available via the Python Package Index</u>) is a Python wrapper for the GETS R package (<u>available via CRAN</u>). All our methods and underlying code are openly available in a dedicated <u>OpenPrescribing GitHub repository</u>. The full results, summary statistics of changes detected and top 10 CCGs for the ranked change can be seen in the <u>full CCG method notebook</u> and the full results, summary statistics of changes detected and top 10 practices for the ranked change can be seen in the <u>full practice method notebook</u>. All organisations that existed in the study period (including those that have since closed or been replaced) are included in these reports.

## Results

We identified substantial reductions in at least 49% of all CCGs and practices for all measures; summary statistics for these reductions are provided in Table 1. Note that these data describe *all* substantial reductions detected, i.e., before filtering for a top 20% starting value. For both CCGs and practices, reductions are on average greater for both high-dose opioid prescribing measures as compared to those observed for the total OME measure, though the IQR values demonstrate that there is also more variability in the high-dose opioid prescribing measures. Reductions appear more modest amongst CCGs than practices (with lower medians and lower maximum values) but these reductions may be more consistent (with lower variability and greater minimum values observed in CCGs as compared to practices). There is at least one practice in each measure where the reduction is almost 99-100% and at least one practice where the reduction detected is very close to zero.

**Table 1:** Summary of all opioid reductions identified using the change detection algorithm. Count indicates the number of organisations (CCGs or practices) in which a reduction was identified. Median, IQR, Min and Max summarise the size of the reductions identified in those organisations (expressed as % reduction from the pre-drop value to the end-drop value).

| Measure | Count | Median | IQR | Min | Max |
| --- | --- | --- | --- | --- | --- |
| **Clinical commissioning groups (n=191)** |  |  |  |  |  |
| Total OME | 94 | 15.1 | 6.9 | 9.0 | 32.8 |
| High dose opioids as percentage regular opioids | 168 | 19.0 | 12.0 | 3.6 | 41.5 |
| High dose opioids per 1000 patients | 115 | 22.2 | 12.8 | 1.0 | 45.4 |
| **Practices (n=7460)** |  |  |  |  |  |
| Total OME | 4100 | 28.2 | 19.8 | 0.1 | 99.1 |
| High dose opioids as percentage regular opioids | 4632 | 47.7 | 32.9 | 0.0 | 100.0 |
| High dose opioids per 1000 patients | 4334 | 56.0 | 35.7 | 0.0 | 100.0 |

### Clinical Commissioning Groups

Table 2 illustrates the CCGs who exhibited the biggest reduction in each of the three OpenPrescribing measures over the study period, detailing the proportion of change and the month in which the change started. Note that these CCGs meet the criteria for identification, i.e., their prescribing rate immediately before the reduction was in the top 20% of all CCGs.

**Table 2:**
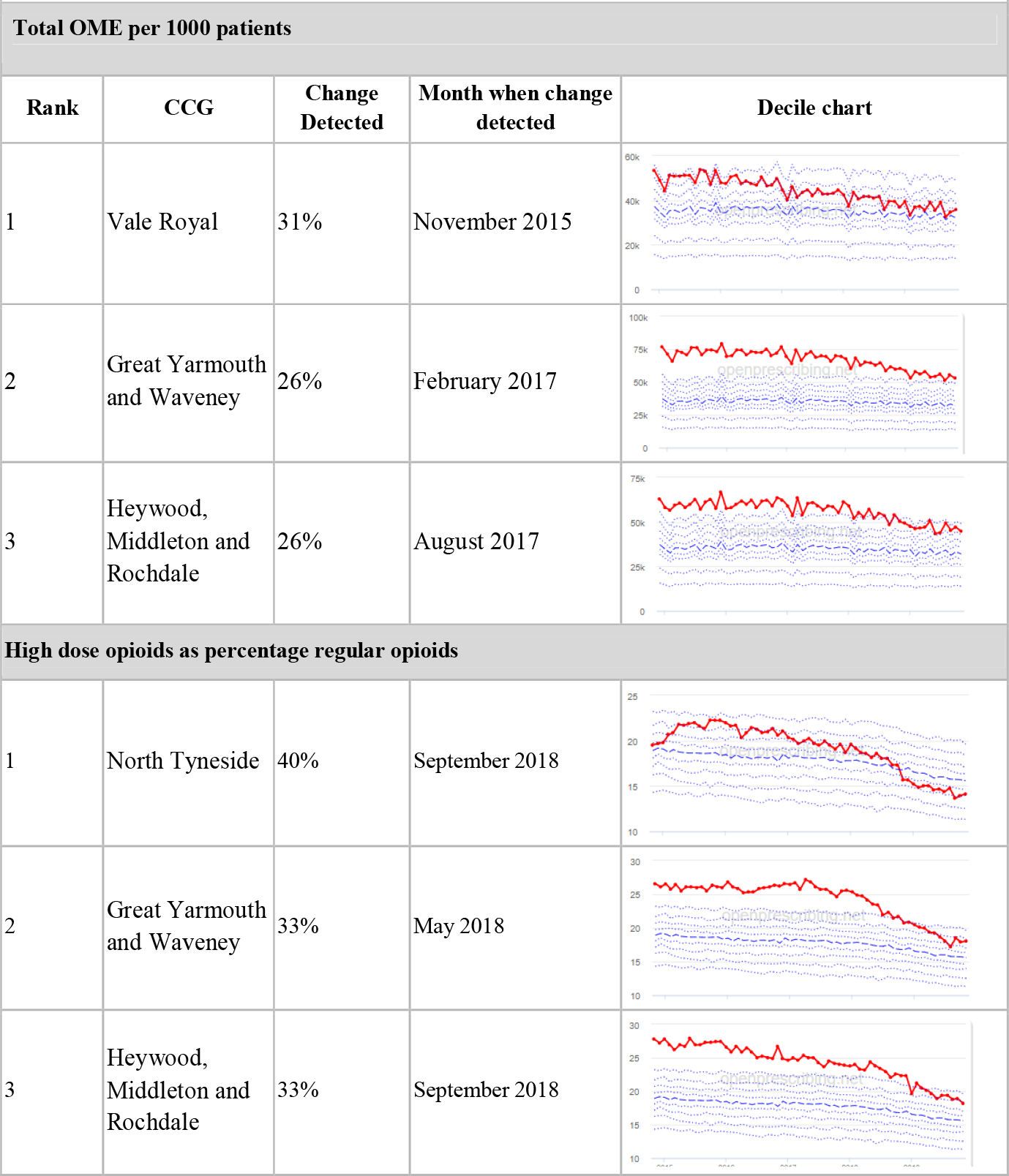
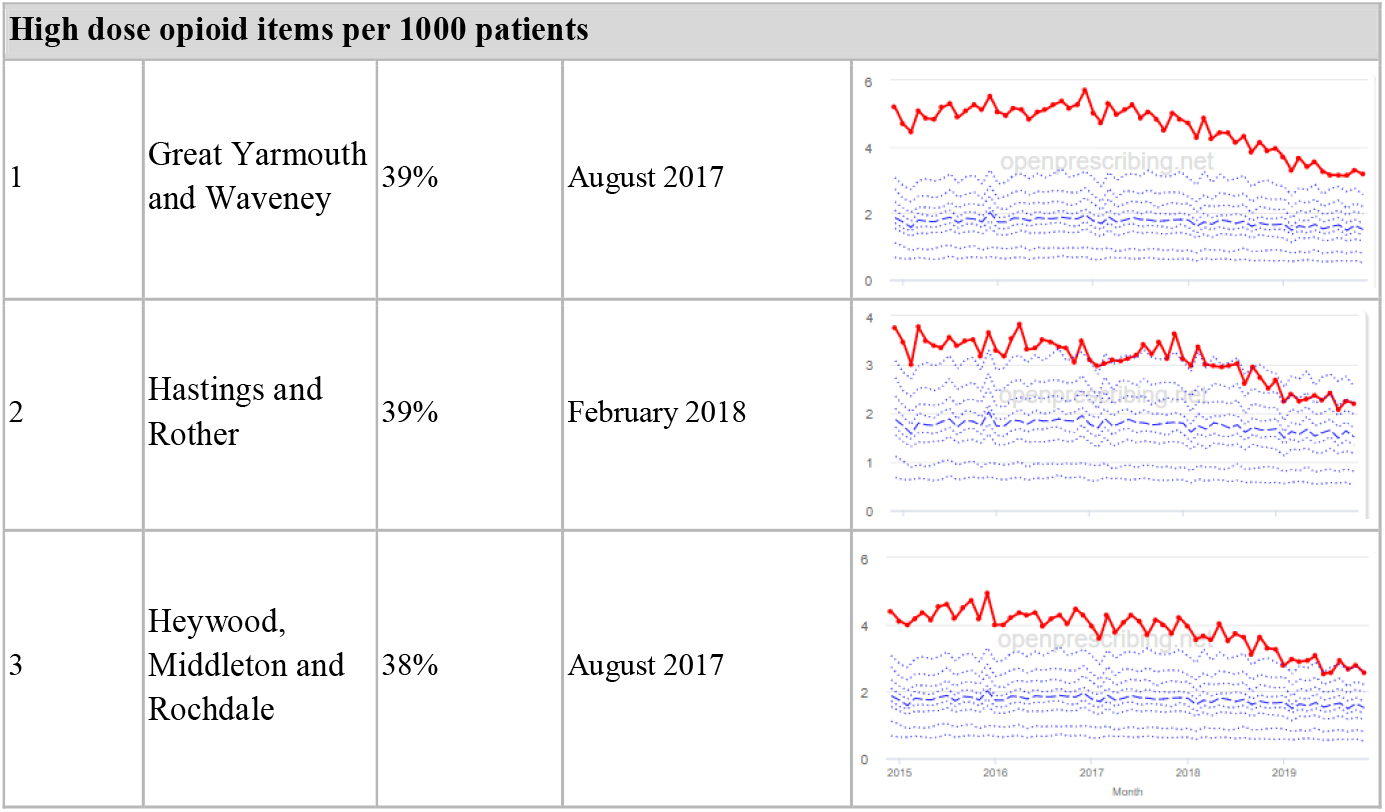
Ranked top 3 CCGs exhibiting a reduction in each of the OpenPrescribing Opioid Measures (December 2014 - November 2019). The decile chart shows the prescription rate for the CCG as a thick red line; prescribing rates for all other CCGs are summarised using deciles (dotted blue lines) with the median highlighted (thick dashed blue line).

The total OME opioid, the measure most sensitive to overall reductions in opioid prescribing, shows gradual reduction over time in all three CCGs, with the algorithm identifying a reduction of up to 31%. The results for the two regular high-dose opioid measures also exhibit a gradual reduction over time but appear to capture greater reductions in regular high-dose opioid prescription, with 40% and 39% reductions identified as a proportion of all regular opioids and per 1000 patients respectively. The trajectory of the deciles for each measure suggest that these reductions are not representative of CCGs overall.

### Practices

Table 3 illustrates the practices who exhibited the biggest change in each of the three OpenPrescribing measures over the study period, detailing the proportion of change and the month in which the change started. Note that these practices meet the criteria for identification as described in the “Statistical Methods” section, i.e., their prescribing rate immediately before the reduction was in the top 20% of all practices.

**Table 3:**
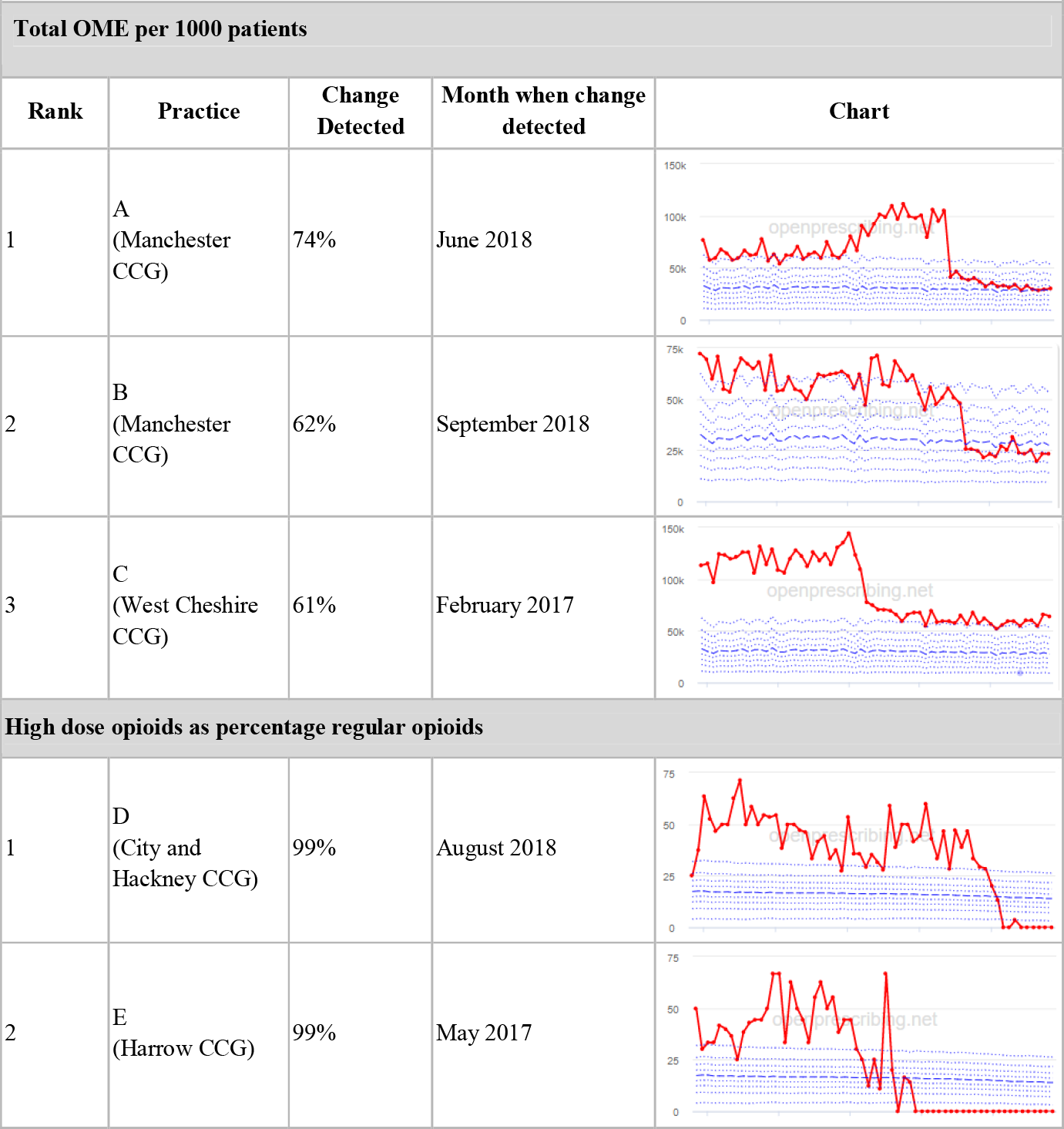
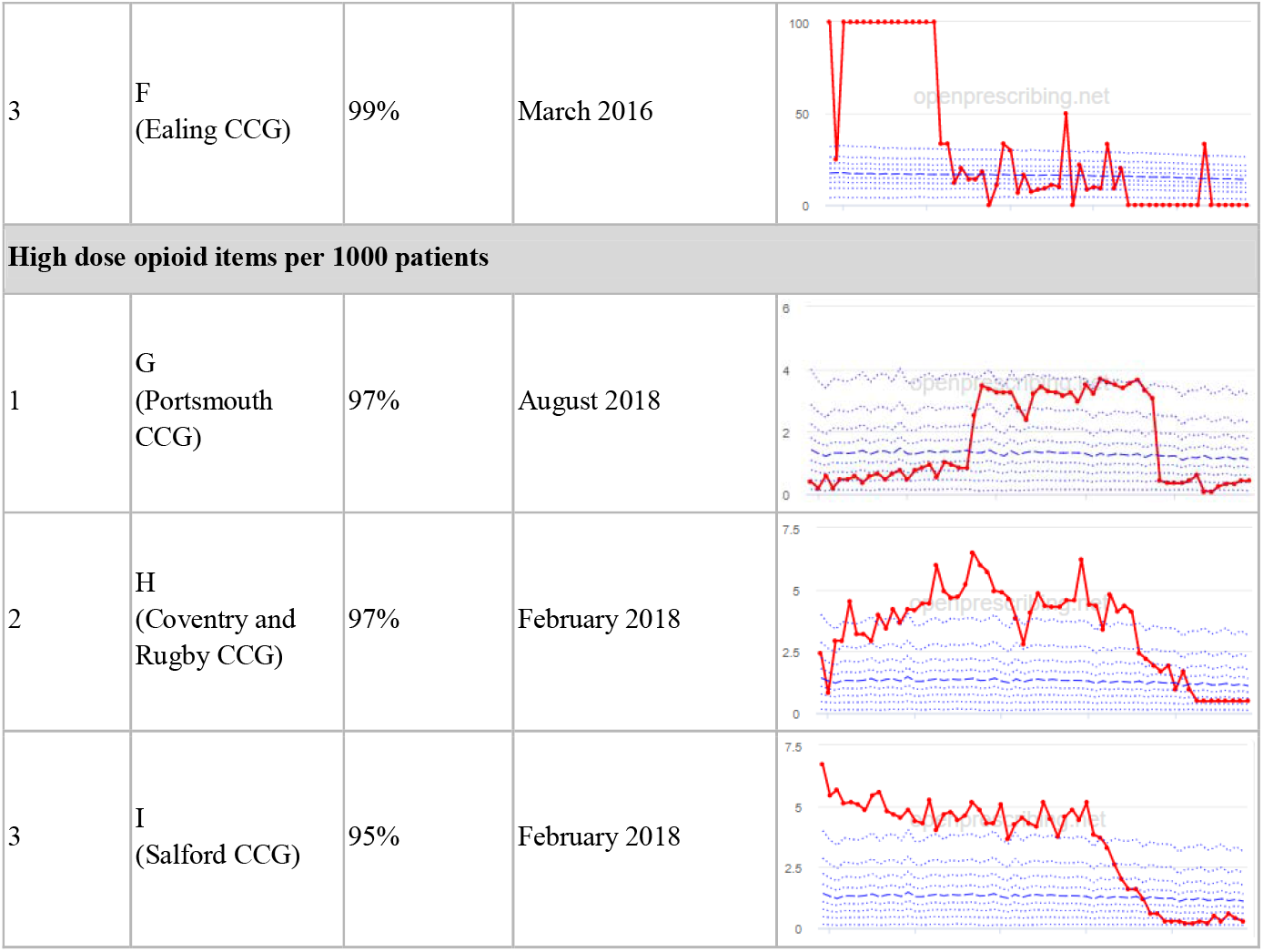
Ranked top 3 practices exhibiting a reduction in each of the OpenPrescribing Opioid Measures (December 2014 - November 2019). The decile chart shows the prescription rate for the practice as a thick red line; prescribing rates for all other practices are summarised using deciles (dotted blue lines) with the median highlighted (thick dashed blue line).

|  |  |  |  |  |
| --- | --- | --- | --- | --- |
| **Total OME per 1000 patients** |  |  |  |  |
| **Rank** | **Practice** | **Change Detected** | **Month when change detected** | **Chart** |
| 1 | A<br>(Manchester CCG) | 74% | June 2018 |  |
| 2 | B<br>(Manchester CCG) | 62% | September 2018 |  |
| 3 | C<br>(West Cheshire CCG) | 61% | February 2017 |  |
| **High dose opioids as percentage regular opioids** |  |  |  |  |
| 1 | D<br>(City and Hackney CCG) | 99% | August 2018 |  |
| 2 | E<br>(Harrow CCG) | 99% | May 2017 |  |
| 3 | F<br>(Ealing CCG) | 99% | March 2016 |  |
| **High dose opioid items per 1000 patients** |  |  |  |  |
| 1 | G<br>(Portsmouth CCG) | 97% | August 2018 |  |
| 2 | H<br>(Coventry and Rugby CCG) | 97% | February 2018 |  |
| 3 | I<br>(Salford CCG) | 95% | February 2018 |  |

The practice time series (Table 3) are noticeably different to those of the CCGs (Table 2): dramatic, short-term changes are common in these practices and change magnitudes are much larger. In the case of the regular high-dose opioids as a percentage of all opioids, all three practices are seen to completely eliminate all regular high-dose opioids for several months; similarly, very low values are observed for the top three practices with regards to reductions in high dose opioid items per 1000 patients. Again, the trajectory of the deciles for each measure suggest that these reductions are not representative of practices overall.

## Discussion

### Summary

Using a data-driven approach we have identified, in over 7,000 practices across 191 CCGs in England, significant reductions in three measures of opioid prescribing (Table 1). These organisations have then been ranked by magnitude of reduction to identify where the largest reductions have been realised. The top ranked CCGs exhibit slow and gradual reduction in opioid use (Tables 1, 2); by contrast the top ranked practices exhibit rapid and sudden reductions over a few months (Tables 1, 3). Opioid prescribing and treatment of pain more broadly can be complex but our findings illustrate that some CCGs and practices appear to have significantly reduced their prescribing of opioids over the study period, more so than many of their peers.

### Strengths and limitations

This analysis has taken into account most (87.7%, 6543 of 7458) typical primary care practices in England, thereby minimising the risk of biased sampling. Executing this analysis in an existing, open platform like OpenPrescribing ensures accountability and transparency—both identified as priorities in the PHE report (9)—by default: all code in this study, from data curation to completed output, is shared openly on GitHub and the Python Package Index. Furthermore, there exists a robust and tested framework with which relevant new measures can be introduced or existing measures can be amended as required in order to respond to any evolving change in tackling opioid dependency and abuse. Our use of OME conversion permits the reporting of trends for opioid medicines overall, whilst accounting for variation in strength.

We also note some limitations. Firstly, the NHSBSA dataset does not include secondary care prescriptions as this was unavailable at the time of the study (17) and as such the opioid measures implemented here may underestimate the extent of opioid prescribing nationally, although financial data would indicate that the vast majority of analgesics (BNF Section 4.7 which includes the BNF Subsection 4.7.2 Opioid analgesics) are prescribed in primary care (18). Secondly, we acknowledge that practice-level time series data in particular could be significantly impacted by local circumstances, including low patient numbers, a change in patient population, a change to prescription frequency (e.g., from weekly to monthly scripts), or a shift in responsibility of opioid prescribing (e.g., from primary to secondary care); and therefore that an apparent reduction in any opioid measure may not be due to a successful intervention. For example, practice G (Table 3) rapidly increased their high dose opioid items per 1000 patients in 2016 followed by a similar rapid reduction two years later; this could be due to a change to daily prescribing as can be clinically justified for some patients.

While we acknowledge these limitations, It is important to note that the intention of this methodology was always to rank or prioritise organisations for further investigation, rather than definitively ascribe reductions in opioid prescribing to successful interventions.

### Findings in context

The PHE review identified evidence of tentative progress in reducing opioid prescribing between 2015/16 and 2017/18 (9). Our analysis includes and extends this time period, and finds evidence that some organisations may be driving this tentative progress more than others (e.g., the CCGs reported in Table 2).

We do have evidence that one of the organisations that has emerged as a potential candidate by our methodology is a genuine example of improved performance. Between 2017 and 2019, Great Yarmouth and Waveney designed and implemented an extensive programme of opioid reduction interventions, including target trajectories for improvement; incentive schemes for clinicians; dialogue with practice pharmacists, patient groups and relevant clinical groups (e.g., prescribing leads and pain management teams); new patient information materials; collecting case studies for discussion; and associated press and social media to raise awareness. While this CCG still suffers from high levels of opioid prescribing, rates have reduced significantly, with the organisation being recognised for this progress nationally (19). Our methodology ranked Great Yarmouth and Waveney as 1st (reduction of 39% starting in August 2017) for the high-dose opioid prescribing per 1000 patients and 2nd (reduction of 33% starting in May 2018) for the high-dose opioid prescribing as percentage regular opioid prescribing, aligning with the period of intervention implementation.

### Implications for research and policy

We are seeking to implement this methodology as a new “Improvement Radar” tool on OpenPrescribing, with the intention of systematically identifying candidates for further qualitative research across multiple important public health prescribing measures. It is our experience that best practice is typically defined by organisations identifying themselves as having improved, following implementation and internal assessment of interventions. Using the Improvement Radar, policymakers interested in spreading best practice can systematically identify organisations who may have already implemented effective interventions. However it is critical that policymakers undertake further investigations for reasons outlined in the limitations. This tool offers an opportunity to reduce resources needed to identify best practice. Similarly local medicines optimisation (MO) teams may wish to use data and tools like this to identify peers across the country who have already delivered successful interventions to inform local initiatives.

## Conclusions

We have demonstrated that data-driven approaches to detect substantial changes in time series data have potential value in the context of opioid prescribing. We have been able to rank organisations with regards to the extent of opioid prescribing reduction; organisations occupying the top of that list show large drops which warrant further qualitative investigation and could be indicative of success in tackling an important public health concern.

Should this further qualitative research reveal that reductions have been driven by well designed and well implemented interventions, methods of best practice will have been identified using an unbiased, evidence-based approach. The organisations found to be implementing this best practice may have valuable insights, approaches and policies to share regarding how positive change can be achieved elsewhere. It also demonstrates, particularly in the most robust and gradual change observed amongst CCGs, that positive change is possible, and therefore that continued and wider success in reducing opioid prescribing is dependent, at least in part, in closing the implementation gap.

## Data Availability

All data produced are available online at https://github.com/ebmdatalab/opioids-change-detection-notebook

## List of abbreviations

CCG: clinical commissioning groups
NHSBSA: NHS Business Services Authority
OME: oral morphine equivalence
PHE: Public Health England

## Declarations

### Ethics approval and consent to participate

This study uses open, publicly available data and data that are publicly available from NHS Digital on request; therefore, no ethical approval was required.

### Consent for publication

All authors have given consent for publication.

### Availability of data and materials

The data used for this study are available via the NHSBSA (15). All our methods and underlying code are openly available in a dedicated <u>OpenPrescribing GitHub repository</u>.

### Authors’ contributions

**Conceptualization**: A.J.W., H.J.C., B.G., and B.M.

**Data curation**: P.I., D.E., and S.B.

**Formal analysis**: L.E.M.H., A.J.W., and B.M.

**Funding acquisition**: A.J.W., H.J.C., and B.G.

**Investigation**: L.E.M.H., A.J.W., H.J.C., R.C., and B.M.

**Methodology**: L.E.M.H., A.J.W., H.J.C., R.C., and B.M.

**Resources**: P.I., D.E., and S.B.

**Software**: F.P., P.I., D.E., and S.B.

**Supervision**: B.G.

**Visualization**: L.E.M.H., A.J.W., H.J.C., and B.M.

**Writing - original draft**: L.E.M.H., A.J.W., H.J.C., and B.M.

**Writing - review & editing**: all authors

## Acknowledgements

We are grateful to wider NHS colleagues for discussions that have informed our work on this topic.

